# Applying chemical reaction transition theory to predict the latent transmission dynamics of coronavirus outbreak in China

**DOI:** 10.1101/2020.02.22.20026815

**Authors:** Peng Xu

**Author notes:** Corresponding author; E-mail address (PX).

## Abstract

The recent outbreak of the Covid-19 suggests a rather long latent phase that precludes public health officials to predict the pandemic transmission on time. Here we apply mass action laws and chemical transition theory to propose a kinetic model that accounts for viral transmission dynamics at the latent phase. This model is useful for authorities to make early preventions and control measurements that stop the spread of a deadly new virus.

## Introduction

Starting at the end of 2019, the outbreak of a new coronavirus has caused hundreds of infections in a central city of Hubei Province in China. The mega city (Wuhan) is designed as a major high-speed railway transportation hub with a population of 15 million civilians. Due to the large migration of people to celebrate the Chinese lunar New year (also called Spring Festival, Jan 25 of 2020), the virus spreads rapidly in Wuhan and neighboring cities. This new coronavirus (2019-nCoV) could cause severe acute respiratory infections (SARIs) with lung damage [1]. The early onset of the SARIs was initially characterized as ‘non-transmissible’ among humans by the local government and health specialist. But later was corrected to ‘transmissible’ among humans. Public health authorities and expert flew to Wuhan and announced a first-grade infectious disease alarm: major railways, airport and bus stations were locked down to reduce every possible chance of virus transmission. Tens of hundreds of people were suspected to be infected by this virus and require hospitalization and special health care in Wuhan. However, large populations of the local patient could not be hospitalized due to the limited medical resources, worsening the pandemics at the worst possible time of the year (Spring Festival). The entire nation was in panic and worry about the spread of the virus; populations with pneumonia-like symptoms or Wuhan travel history were quarantined, flights and railway transportation were suspended around the nation, and new health centers specialized in infectious disease were constructed to accommodate the daily-increasing patients.

Up to date, epidemiologists and health care professionals have used a number of mathematical models to predict the consequence and infection trajectories of the virus outbreak. Both kinetics [2, 3] and statistics-based [4, 5] models have predicted the basic reproduction number ranging from 2.0 to 4.0, which signify an alarming sign to designate this new virus as ‘highly transmissible and communicable’. In the past few days, the population of the infected people increased dramatically in Wuhan and other cities of China, due to the relative long latent period of this new virus. Latency period is defined as the time between infection with a virus and the appearance of symptoms. The accurate estimation of the latency period determines which kind of control measurement actions the health officials and governments should take. Latency period can also be used to predict the population of the people who is exposed to the virus but not yet converted to diseased state. The dormant virus is still active and transmissible, which is a hidden factor that brings about the most unpredictability and uncertainty of the current models.

### Chemical transition model development

Latency period is the transition from ‘exposed state’ to ‘diseased state’, which is the rate-limiting step of the disease transmission process. In the fundamental elements of chemical reaction theory, chemical transition state is generally applicable to most of radicals or intermediate complex-associated reactions, including examples of photoactivation reaction or enzyme catalysis. Because of the synergy between the latent period of the ‘exposed population’ and the chemical transition of complex reactions, we here propose a viral transmission model that mimics the encounter complex state of virus to predict the exposed population, which we termed as SM* (Fig. 1), to describe the ‘exposed population’ at the latent period. The active virus source M* can be released from both the infected (I) and the exposed population (SM*), but not the deceased population D (assuming complete containment of this category). A simplified 7-step transmission mechanism has been exemplified in Fig. 1. Step 1 describes how fast a normal population could be converted to susceptible population who is at risk of infection. Step 2 describes the virus-releasing kinetics of infected people, with each infected people releasing *n* virus or virus-contaminated subjects (i.e. aerosols). Step 3 describes the infection kinetics of the susceptible people that is converted to an ‘exposed population’ at the latent phase. A reversible mechanism is proposed here since some of the exposed population didn’t develop symptoms and could naturally return to the susceptible state. Step 4 describes the infection kinetics of the exposed population that is converted to infectious state. Step 5 and step 6 describe the recovery and death kinetics from the infectious population, respectively. Step 7 describes the decay kinetics of the active viral carrier to an inactive viral carrier. With fundamental mass action laws, we have formulated eight differential equations (Eqn1 to Eqn8) to describe the viral transmission kinetics based on the 7-step infectious mechanism. The mass balance of total populations (Ntot) and the total viral transmissible media (Mtot) were conseved by Eqn. 9 and Eqn. 10, respectively.

**Fig. 1.**
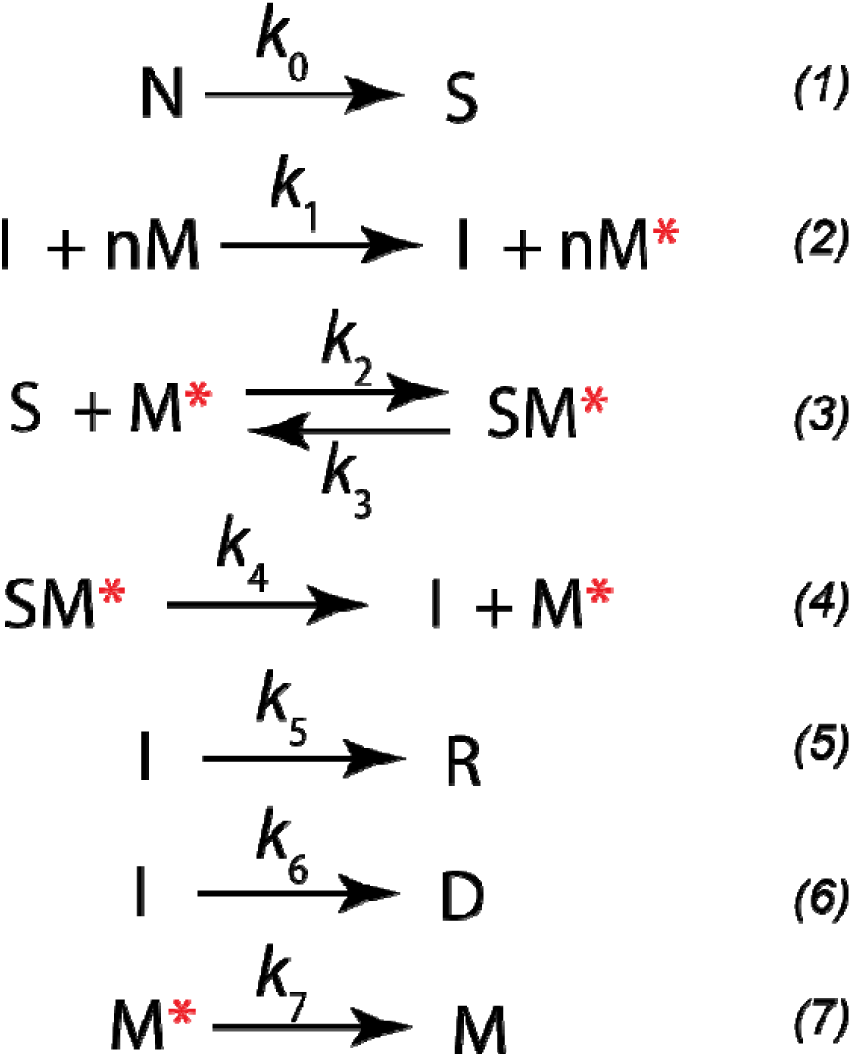
Chemical transition theory to predict the transmission of coronavirus. N: normal health population; S: susceptible population; M: inactive media that may carry virus; M*: active viral source; SM*: exposed population that carry the active virus (Latent state); R: recovered population; D: deceased population.

### Latent transmission dynamics prediction

With the recently reported cases of infection, death rate and exposed population, we used a set of parameters to simulate the model and predict the pandemic trajectory. In particular, our model is able to predict the trend of exposed population who is in the latent phase, and the total number of active viral source that remain infectious in the community (Fig. 2). We also introduced a migration rate (*alpha* = 0.1 people/week, which is unrealistically small in the current model) to describe potential flow of people moving into the system. With about 0.83 weeks of latency period (*1/k*_4_, *where k*_4_ is 1.20/week in Fig. 2), we predict that the peak latent population is about 56,100 cases on Feb. 20^th^ (0.5 week), and peak population of infected people is about 91,000 cases around March. 10^th^ (3.3 weeks). Around March 15^th^ (4 weeks), the number of active virial carrier start declining (Fig. 2) and 57.7% of the infected population will be cured or recovered. A projected deceased population is about 3,750 at a time window of 20 weeks (on June 5^th^).

**Fig. 2.**
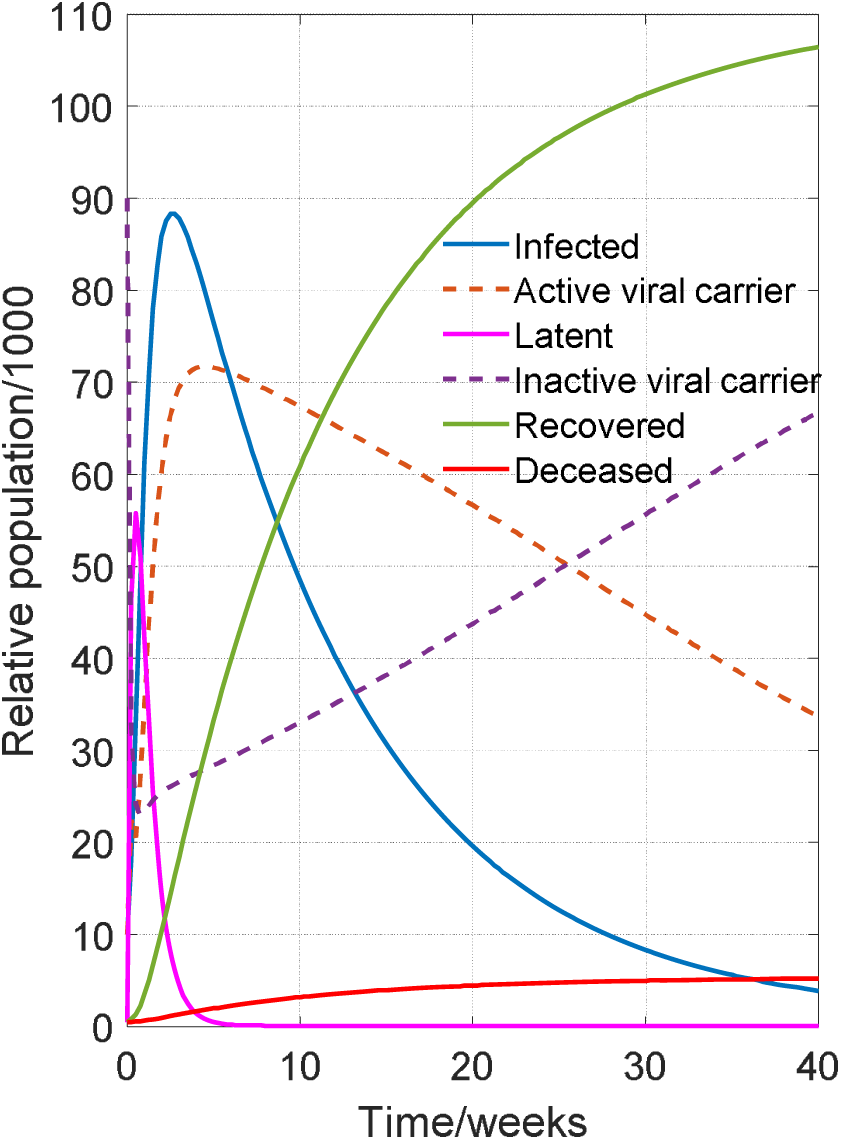
Representative population dynamics of infected, latent, recovered and deceased states predicted by the proposed model. Active viral carrier describes the number of active viral source released from the infected and latent population.

By considering the latency period factor, this model is also useful to predict the dynamic trajectory of the population at the latent phase (Fig. 3). The latent population with a 10-week latency period (88,500 cases) is about 75.2% higher than the latent population (about 50,500 cases) with a 0.67-week latency period; as a result (Fig. 3). With a 10-week latency period, the latent population (88,500 cases) is about 2.1-fold than the infected population (42,000 cases). However, the peak population of the infected people (91,000 cases) is 80.2% higher than the latent population (about 50,500 cases) with a 0.67-week latency period (Fig. 3). This result indicates that a rather long latency period decreases the total number of infected people, and the peak infection population is also postponed (Fig. 3). This prediction suggests that we must have an accurate estimation of the latency period before any coercive or preventative actions be taken. Based on the current report from different regions in China, it appears that some of the exposed people haven’t shown any symptoms up to 2∼3 weeks [6], but this latent population will keep spreading the virus and transmitting the disease, a more severe transmission than the SARS pandemic in 2003 [7]. This uncertainty of the latency period complicates the infection measurements and may deteriorate the situation where people thought they have taken enough precautions to prevent the next outbreak of the virus.

**Fig. 3.**
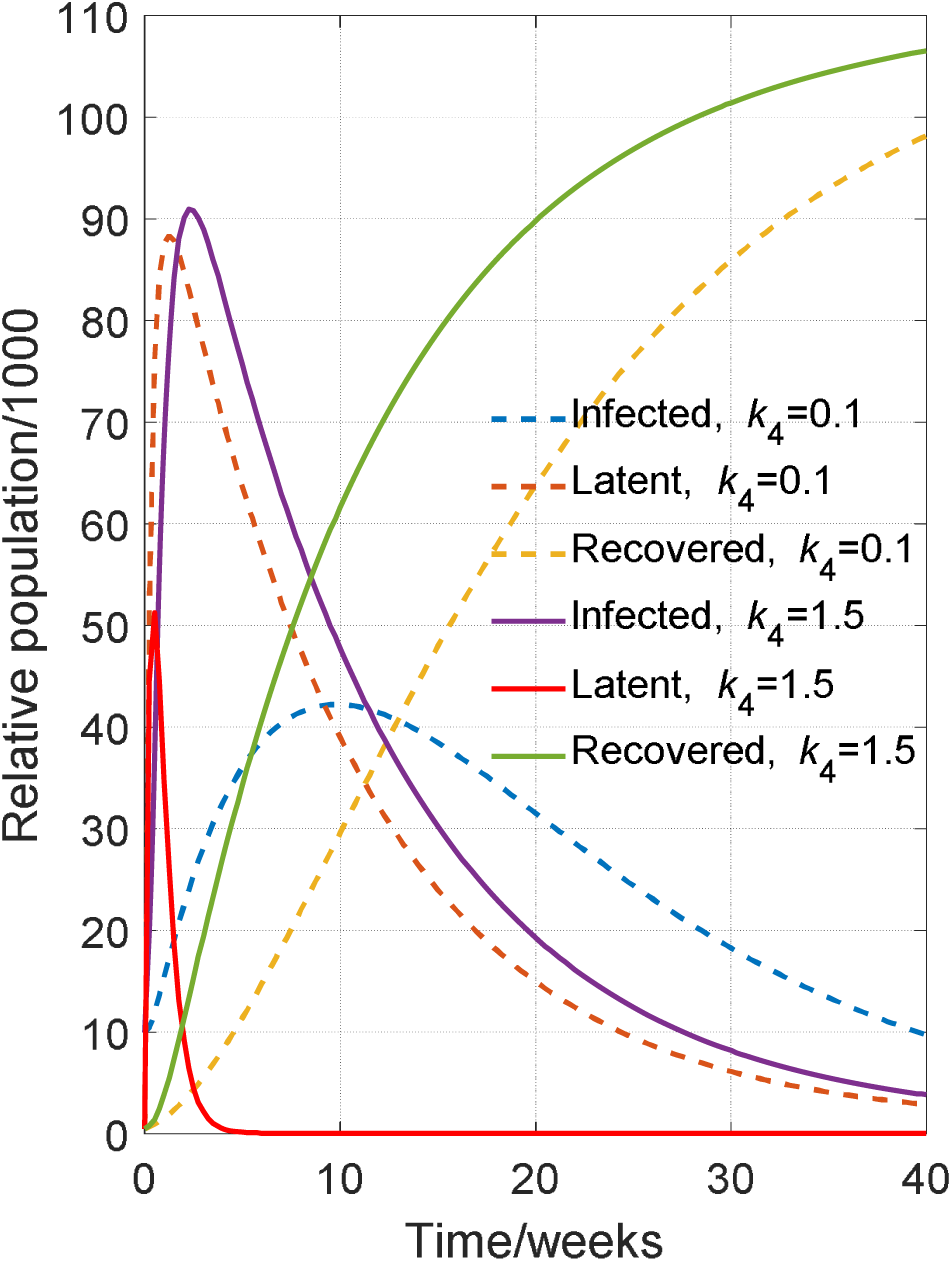
Prediction of the infected, latent and recovered population as a function of the latency period (1/*k*_4_). The dashed line represents a latency period of 10 weeks, and the solid line represents a latency period of 0.67 weeks.

This simplified infectious disease transmission model takes into consideration of the latency period. The accurate prediction of the latency period will be critical for public health officials and authorities to make early preventions and take rigorous control measurements that stop the further spread of this deadly new virus.

### Computational methods

Matlab R2017b was used as the computational platform and installed on a Windows 7 professional operation system with Intel Core i3-6100 CPU processor at speed of 3.70 GHz. The installed memory (RAM) is 4.0 GHz. Matlab symbolic language package coupled with LaTex makeup language is used to derive and output the symbolic equations (Supplementary files). Numerical solutions for the proposed infection model in Fig. 2 were computed by numerical ODE45 solver. Matlab code has been compiled into the supplementary file. Parameters (death rate constant of 0.004 people/week is used) were obtained by estimating the confirmed infection, death and recovered population reported by the officials. Our simulation is run with a constant migrate rate alpha = 0.1 people/week and a total population is confined at 120,000. The model can be scaled up to a real city, if accurate estimation of the kinetic parameter is feasible.

## Data Availability

yes, all data is available

## Acknowledgments

Dr. Xu would like to acknowledge the Bill and Melinda Gates Foundation (OPP1188443) for financially supporting this project.

## Equations used in this study

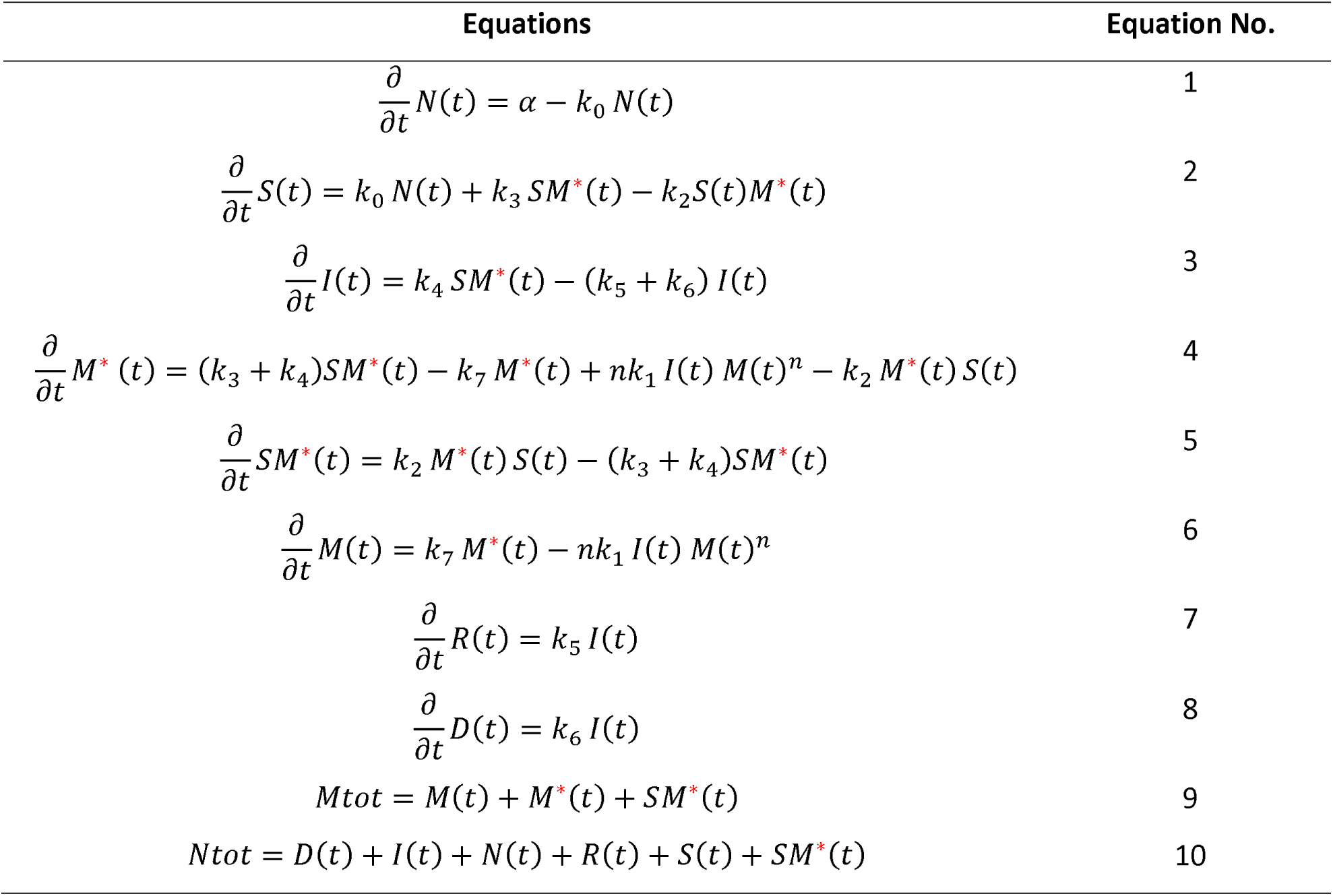

| Equations | Equation No. |
| --- | --- |
| $\frac{\partial}{\partial t} N(t) = \alpha - k_0 N(t)$ | 1 |
| $\frac{\partial}{\partial t} S(t) = k_0 N(t) + k_3 SM^*(t) - k_2 S(t)M^*(t)$ | 2 |
| $\frac{\partial}{\partial t} I(t) = k_4 SM^*(t) - (k_5 + k_6) I(t)$ | 3 |
| $\frac{\partial}{\partial t} M^*(t) = (k_3 + k_4)SM^*(t) - k_7 M^*(t) + nk_1 I(t) M(t)^n - k_2 M^*(t) S(t)$ | 4 |
| $\frac{\partial}{\partial t} SM^*(t) = k_2 M^*(t) S(t) - (k_3 + k_4)SM^*(t)$ | 5 |
| $\frac{\partial}{\partial t} M(t) = k_7 M^*(t) - nk_1 I(t) M(t)^n$ | 6 |
| $\frac{\partial}{\partial t} R(t) = k_5 I(t)$ | 7 |
| $\frac{\partial}{\partial t} D(t) = k_6 I(t)$ | 8 |
| $M_{tot} = M(t) + M^*(t) + SM^*(t)$ | 9 |
| $N_{tot} = D(t) + I(t) + N(t) + R(t) + S(t) + SM^*(t)$ | 10 |

## Symbols used in this study

*α* migration rate of normal population into the system (people/week)

*k*_0_ rate constant of normal population converted to susceptible population (1/week)

*k*_1_ rate constant of infected population releasing the virus (1/week/infected people)

*k*_2_ rate constant of active virus infecting the susceptible population (1/week/virus)

*k*_3_ rate constant of latent population reverted to the susceptible state (1/week)

*k*_4_ rate constant of latent population progressing to the infected state (1/week)

*k*_5_ rate constant of infected population that is recovered (1/week)

*k*_6_ rate constant of infected population that is deceased (1/week)

*k*_7_ rate constant of active virus decaying to the inactive virus (1/week)

*n* virus reproduction number (virus/week)

*N*(*t*) Population at normal state

*S*(*t*) Population at susceptible state

*I*(*t*) Population at infected state

*M*^*^(*t*) Population of active viral carrier

*SM*^*^(*t*) Population at latent phase

*M*(*t*) Population of inactive viral carrier

*R*(*t*) Population that was recovered

*D*(*t*) Population that was deceased

*M*tot Total population of virial carrier

*N*tot Total population in the pool

## Supplementary files

**Appendix 1:**
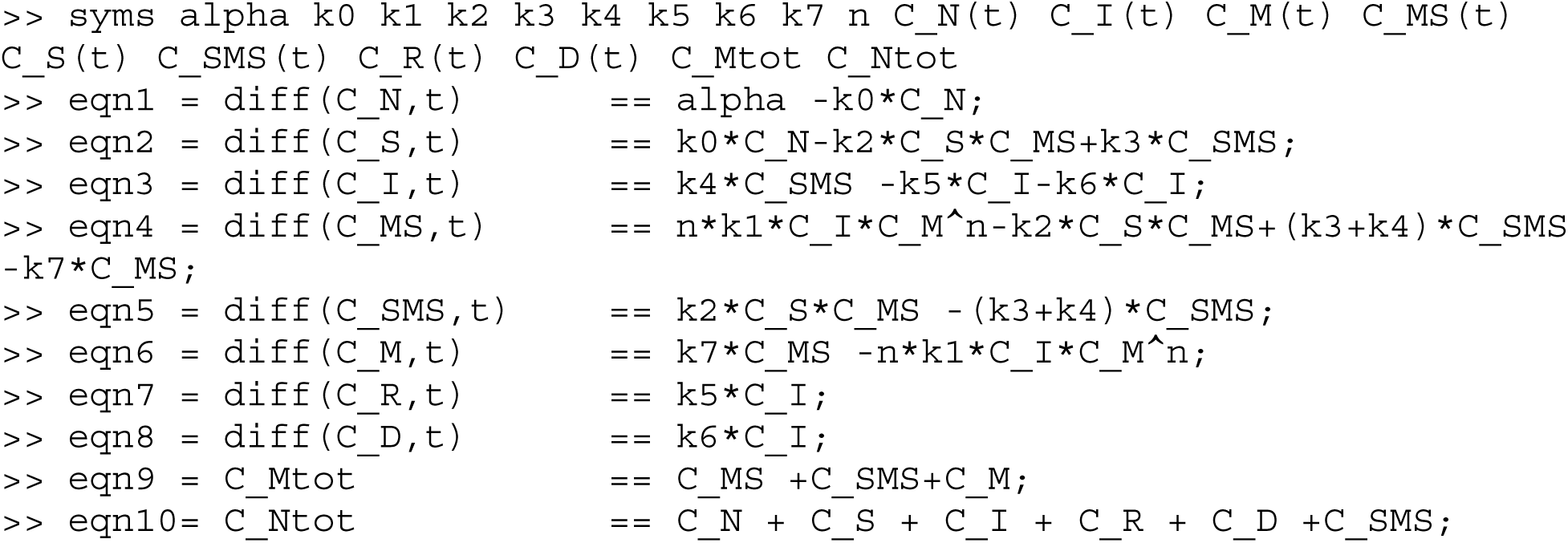
Matlab code to generate the equations used in this work

**Appendix 2:**
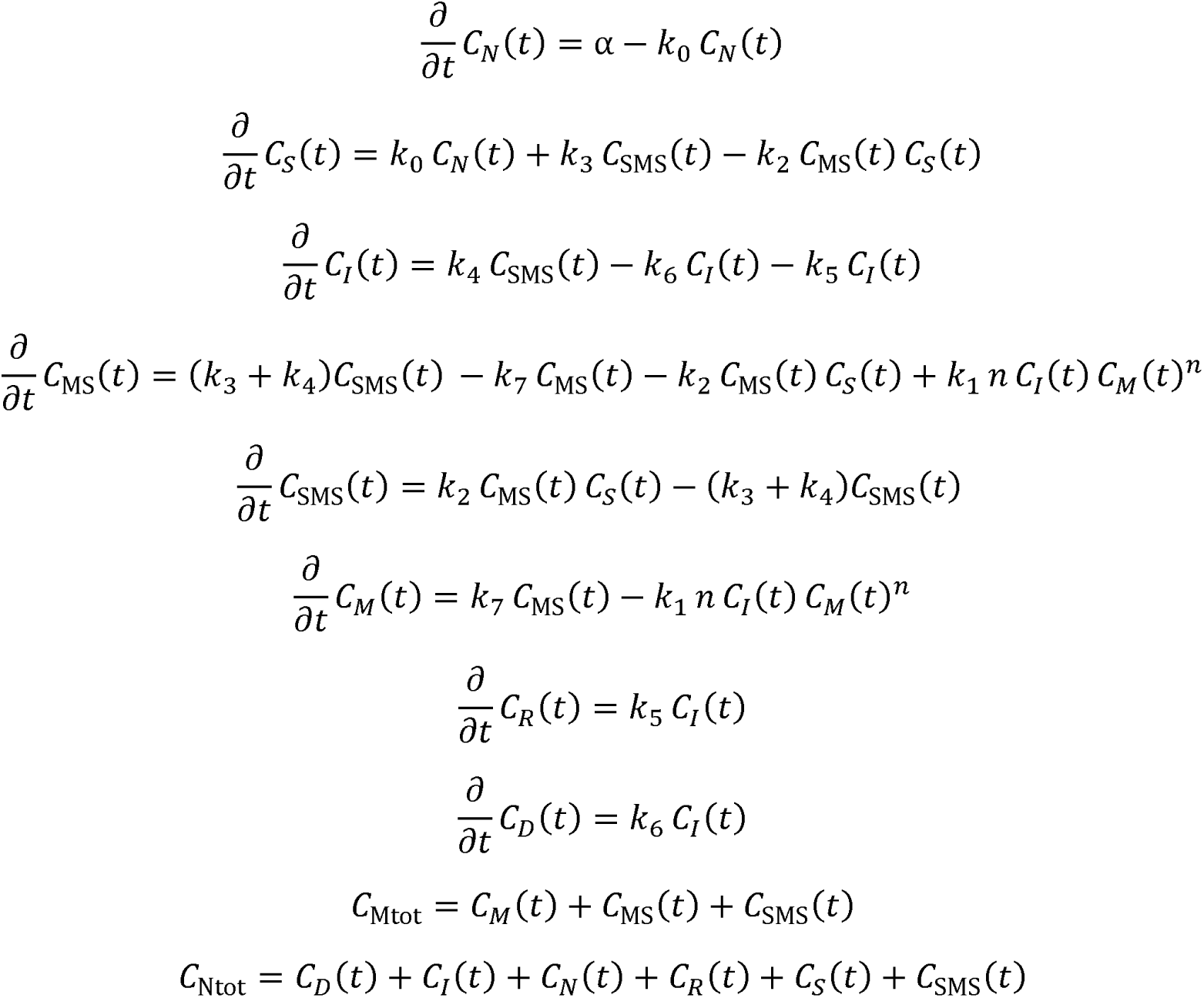
Mass action equations to describe the population change

**Appendix 3:**
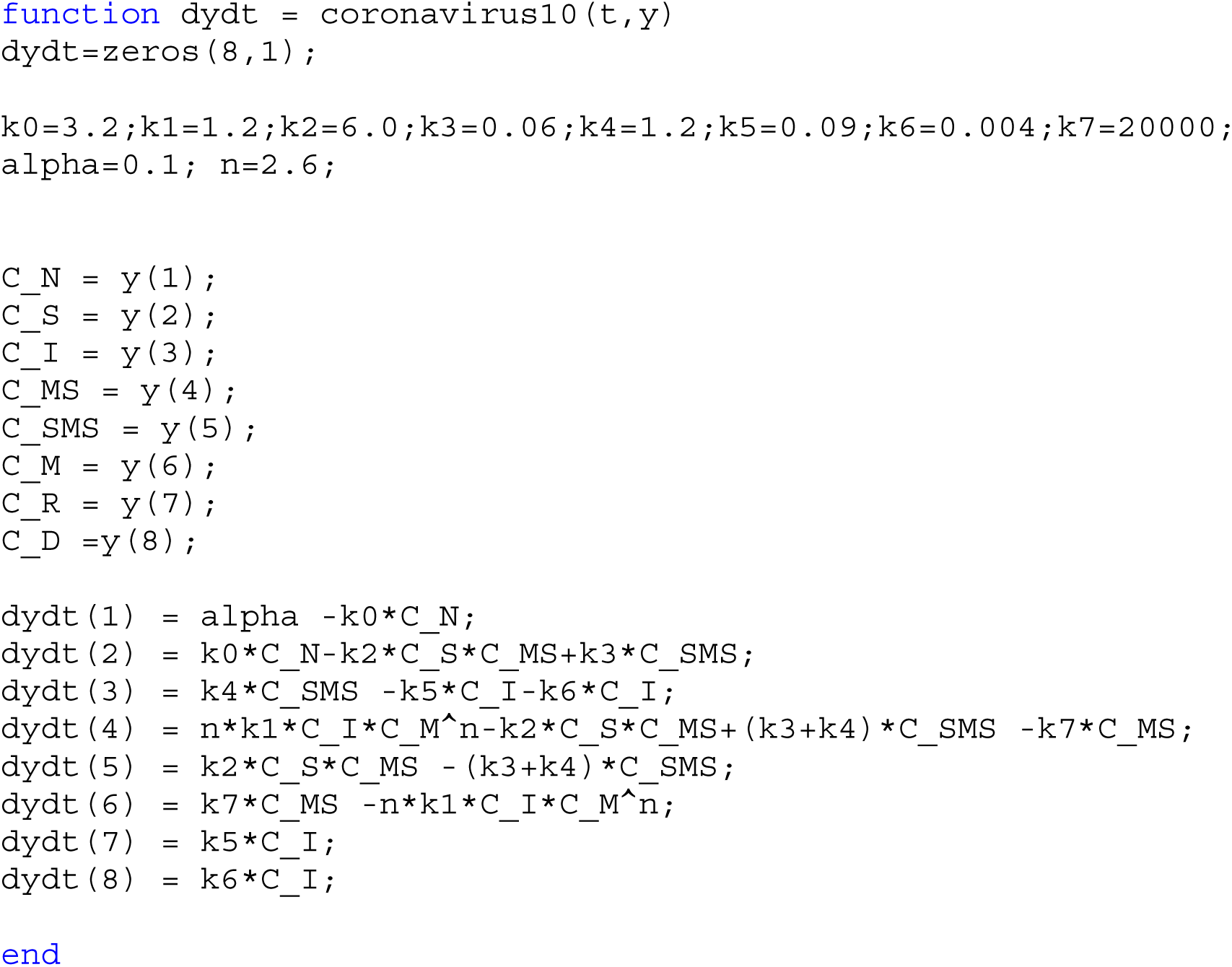
Matlab code to define the differential equations (Eqn1-Eqn8)

**Appendix 4:**
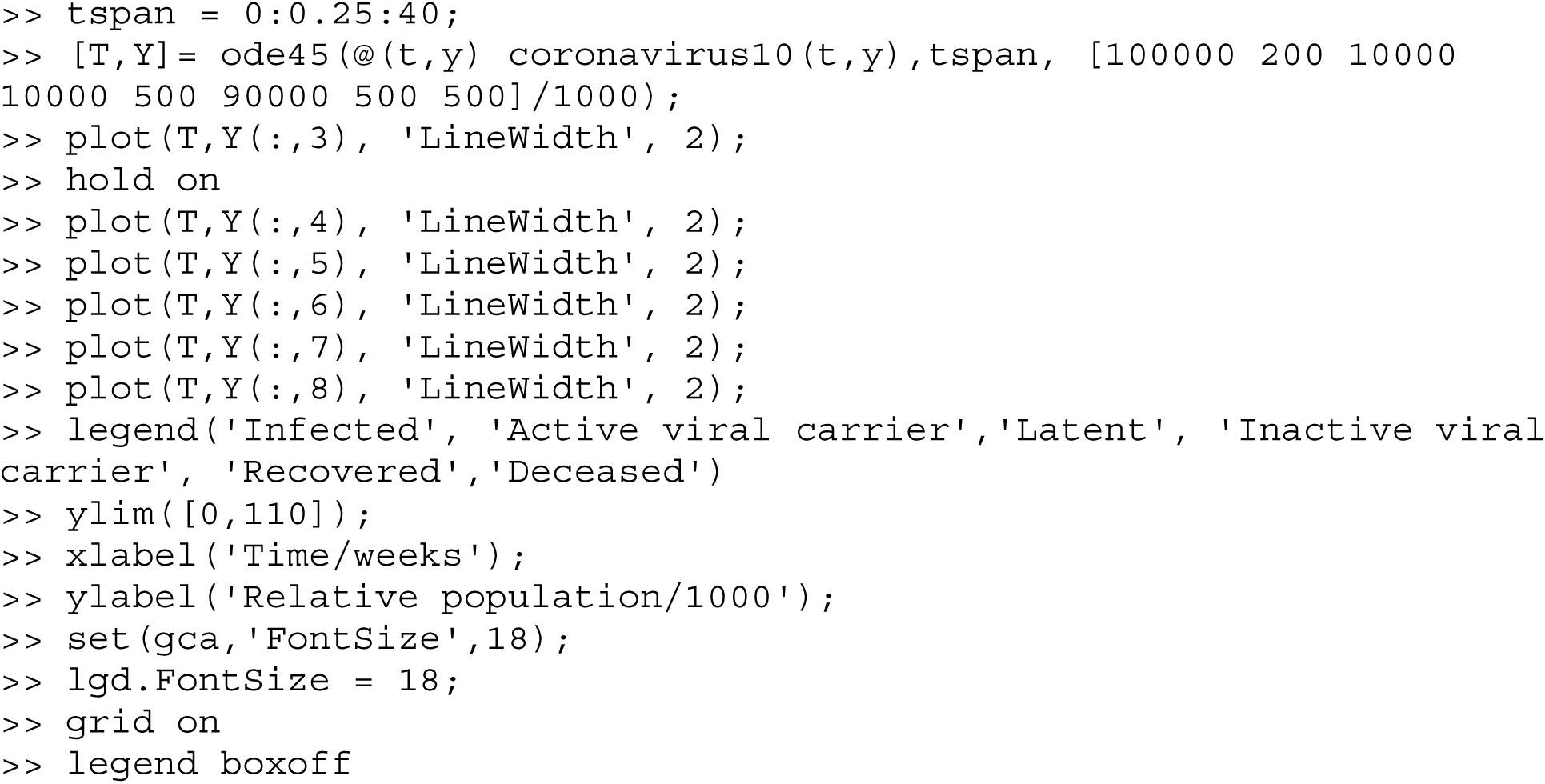
Matlab code to generate Figure 2.

**Appendix 5:**
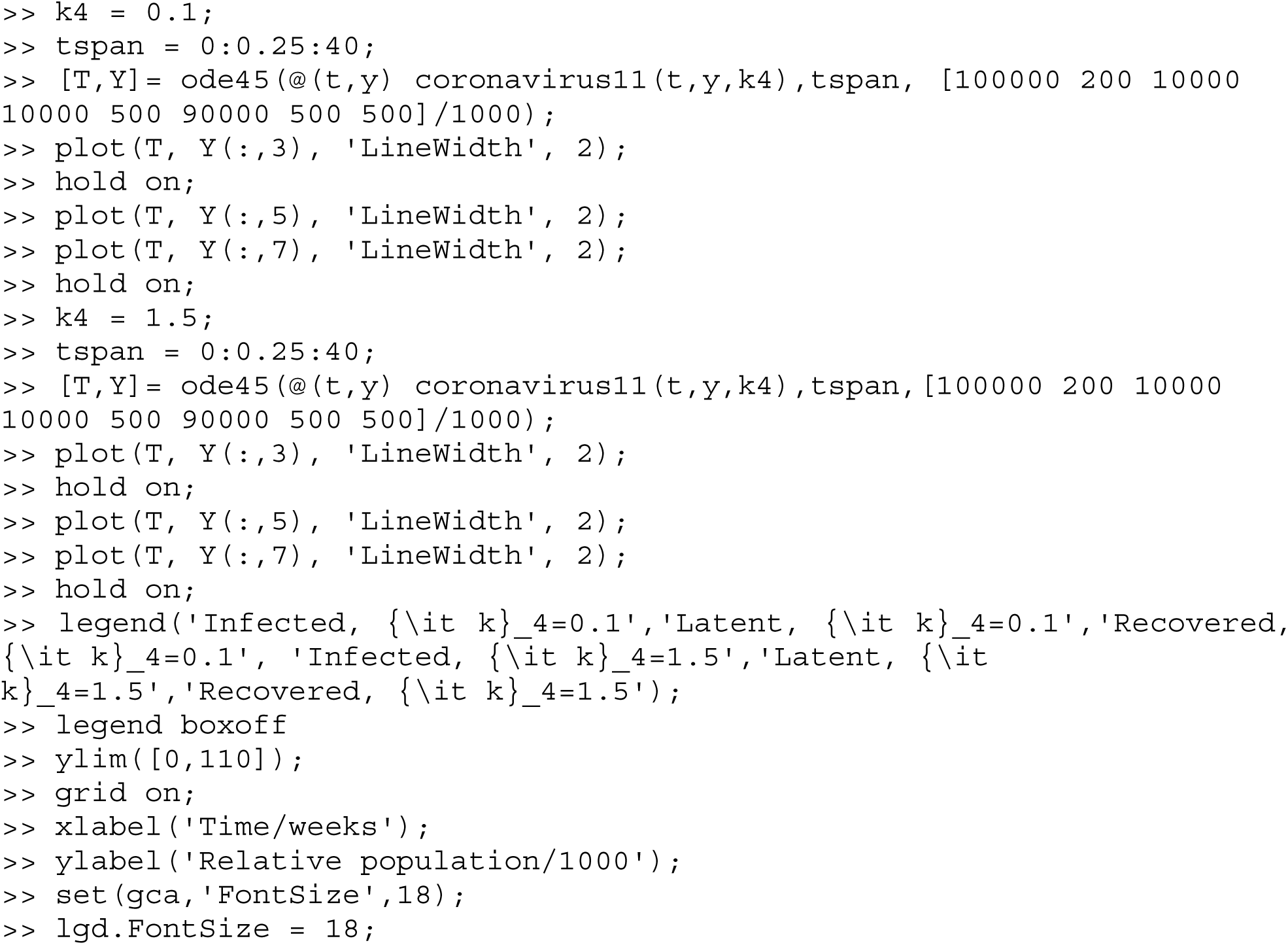
Matlab code to generate Figure 3.

